# Risk Screening in a Medicaid-Managed Pregnancy Medical Home: The Need to Center Maternal Health Outcomes in Public Health Programming

**DOI:** 10.64898/2026.07.16.26358262

**Authors:** Mekhala Dissanayake, Divya Mallampati, Catherine J. Vladutiu, M. Kathryn Menard

**Affiliations:** Department of Epidemiology, University of North Carolina at Chapel Hill, Chapel Hill, NC, USA; Department of Pediatrics, Stanford University, Stanford, CA, USA; Division of Maternal Fetal Medicine, Department of Obstetrics and Gynecology, University of North Carolina at Chapel Hill, Chapel Hill, NC, USA

## Abstract

**Background:** North Carolina Medicaid implemented the Pregnancy Medical Home program to improve access to high-quality maternity care and reduce the risk of adverse perinatal outcomes. Program recipients receive a prenatal risk screening form, originally intended to identify those at high risk of preterm birth and low birth weight, that includes an assessment of social and clinical factors. While prior studies have evaluated whether risk screening can identify pregnancies with higher risk of adverse neonatal outcomes, less is known about the relationship between programmatic risk-stratification and adverse maternal outcomes.

**Objective:** To assess the use of a prenatal risk screen among pregnant Medicaid beneficiaries to identify those at risk of an adverse maternal event.

**Study design:** Linked Medicaid hospital claims, live birth records, and risk screen data from the Pregnancy Medical Home program were used to identify risk factors for adverse maternal events among individuals who gave birth to a liveborn infant in North Carolina between 2014 and 2019. Only those with completed risk screens (75%) were included in the analysis. We used random forest classification to select variables for a multivariable prediction model. We used Poisson regression to model the association between adverse maternal events and selected demographic, psychosocial, clinical, and historical pregnancy characteristics. Adverse maternal events occurring at birth and up to six weeks postpartum included severe maternal morbidity, maternal intensive care unit admission, prolonged birth hospitalization, and postpartum readmissions.

**Results:** A total of 205,916 births met inclusion criteria for this analysis. During the study period, 3.0% of Medicaid beneficiaries had an adverse maternal event occurring between birth and up to six weeks postpartum, including, 0.6% with severe maternal morbidity, 0.9% with an intensive care unit admission at birth, and 1.5% with a prolonged birth hospitalization or postpartum readmission. Maternal age greater than 25 years, Black race, being overweight or obese, smoking, chronic diseases (diabetes, hypertension, mental illness), and pregnancy history characteristics (nulliparity, history of preterm birth, history of hypertensive disorders of pregnancy or gestational diabetes) were associated with an increased risk of adverse maternal events. Modeled together, however, risk factors from the risk form were poorly predictive of the composite outcome. The final model had an Area Under the Curve (AUC) of 0.63 with an optimal sensitivity of 56% and specificity of 63%.

**Conclusion:** Care management during pregnancy is an increasingly relevant topic in public health and prenatal care in the United States. The North Carolina Pregnancy Medical Home is a long-standing and robust Medicaid program that can serve as a model for design and implementation. While this program has effectively designed risk-stratification to identify pregnant people at risk of poor neonatal outcomes who benefit from care management, the risk screen poorly identifies pregnant people at risk of adverse maternal outcomes. Care coordination programs are often designed to optimize neonatal outcomes, and this study highlights the need to center and balance maternal health along with neonatal outcomes to address the needs of a very vulnerable population.

## INTRODUCTION

The maternal mortality rate in the United States is higher than other high-income nations and has not improved in recent decades.^1,2^ Similarly, rates of other adverse maternal health outcomes, such as severe maternal morbidity (SMM) are high in the U.S.^3^ Each year, more than 30,000 women experience these unexpected consequences of labor and delivery that have short- and long-term ramifications for women’s health.^3^ As defined by the Centers for Disease Control and Prevention (CDC), SMM encompasses 21 indicators of conditions and procedures that range in severity and incidence, such as hemorrhage, sepsis, and cardiac arrest, and are considered near-miss events for maternal mortality.^4^ Research identifying drivers of the rise in SMM in the United States have identified rising rates of comorbid conditions, racial disparities, and systems-related factors as contributors.^5^

While SMM is an important outcome to assess in relation to maternal health, it does not encompass all severe patient-centered and systemic outcomes, including those in the postpartum period. SMM does not include intensive care unit (ICU) admissions, hospital readmissions, or prolonged hospitalizations – all of which can occur for reasons not included in the current definition of SMM, such as preeclampsia with severe features and mental health or mood disorders. Importantly, these other indicators of severe maternal outcomes have implications for individuals as well as for the healthcare system.^6,7^

As maternal health and associated health outcomes are complex in the United States, several states and private organizations have implemented models of care that facilitate care management during pregnancy, particularly for the most vulnerable populations.^8–11^ Care management is the effective management of people’s medical, social, and behavioral conditions through a team-based, person- centered approach.^12^ In North Carolina, since 2012, as part of the Pregnancy Medical Home (PMH) Program, Medicaid beneficiaries have been offered screening for clinical and psychosocial risk factors as a way to stratify those at highest risk of preterm birth and identify those who might benefit from care management and additional resources.^13^ Risk screens were typically offered at the first prenatal visit by the provider and answered by the pregnant Medicaid beneficiary. Once identified as being high risk for this adverse neonatal outcome, pregnancy care management through care managers is provided by county health departments. Care managers provide a wide range of services including assistance with adherence to medical appointments, receipt of appropriate patient education or treatment, and resources to meet psychosocial needs. Prior studies on the North Carolina PMH have established that risk screening can identify pregnancies at high risk of preterm birth and low birth weight and that care coordination is associated with reductions in these adverse outcomes.^14,15^ The North Carolina model offers valuable insights into the design of risk screening protocols, risk stratification, the role of care management for diverse populations, and the impact on healthcare utilization and perinatal outcomes.

Pregnancy care management is a growing area of interest to other states, federal, and private entities to foster collaboration among prenatal care providers, social services, public health and other services to support pregnant individuals and improve outcomes. While federal investments in programs that focus on integrated maternal health services are still in their nascent stages,^16^ several state-based programs, like the PMH in North Carolina, have also utilized risk stratification models intended to correlate with neonatal outcomes.^17–19^ It is well-documented that poor neonatal outcomes coincide with adverse maternal outcomes, such as SMM, but there are limited data on whether programmatic risk stratification to predict neonatal outcomes is predictive of poor maternal health outcomes.^20^ The objective of this pragmatic study was to assess whether the use of a risk screen designed to identify those at risk of adverse neonatal outcomes can also identify those at risk of an adverse maternal event in the North Carolina PMH program.

## MATERIALS AND METHODS

This study was a retrospective analysis of live birth records, Medicaid claims, and risk screen data from the PMH program from 2014-2019. These data were linked by Community Care of North Carolina using the maternal Medicaid ID -- 95% of live birth records had a matching birth hospitalization claim, which we required for our analyses (N=319,444). Of these matched records, 228,316 (75%) had risk screen data. Pregnant people were included if they had: 1) risk screen data, 2) were aged 12-55 years at the time of birth; 3) had traditional Medicaid coverage or Medicaid for Pregnant Women; 4) had two or more member months of coverage following birth (i.e., had Medicaid coverage during this time); and 5) had a live birth between January 1, 2014 and December 31, 2019. In the case of a twin or higher order gestation, only one record was retained. This administrative dataset did not capture information on gender identity. Therefore, we use the term pregnant people to be inclusive of all people who are included in our study.

Our primary outcome was a composite of adverse maternal events occurring at birth and up to six weeks postpartum: SMM, maternal ICU admission, and postpartum hospitalizations (prolonged birth hospitalization and postpartum readmissions). We used maternal inpatient Medicaid claims, claims with claim type codes of Inpatient or Medicaid Part A Crossover (Inpatient), to identify each component of our outcome. Consistent with federal definitions and reporting, non-transfusion SMM during the birth hospitalization was defined using International Classification of Diseases (ICD) -9/ ICD-10 diagnosis and procedure codes for the 20 indicators on the birth hospitalization claim.^4^ Maternal ICU admissions were identified using revenue codes.^21^ Following standards published by the Centers for Medicare & Medicaid Services that outlines minimum coverage for hospital stays associated with childbirth (48 hours for vaginal births, and 96 hours for cesarean births),^22^ postpartum hospitalizations were defined using any inpatient claim with dates of service 3-42 days after the infant date of birth for vaginal births and 5-42 days after infant date of birth for cesarean births.

Information on maternal age, race and Hispanic ethnicity, education, rural residence, body mass index (BMI) and plurality were derived from live birth records. The risk screen data provided information on psychosocial characteristics, current clinical factors, and prior pregnancy characteristics. The risk screen form that captures this information has been described in detail elsewhere.^14^ Psychosocial characteristics included: food insecurity, housing instability, history of physical violence, whether the participant had experienced forced sex, substance abuse among partners or family, and the participants’ alcohol or drug use. The current clinical characteristics section included information on multiple gestations, fetal complications, and chronic diseases (diabetes, hypertension, asthma, mental illness, HIV, seizure disorders, renal disease, systematic lupus erythematosus and others), as well as late prenatal care entry and short interpregnancy intervals. The pregnancy history section included information on whether participants had a history of low birth weight, fetal or neonatal death, pregnancy losses, preterm birth, or hypertensive disorders of pregnancy (HDP). We calculated bivariate statistics of each factor and its association with any adverse maternal event.

We used random forest classification to select variables for our prediction model^23,24^ and used the variable importance rankings, which ranks variables by the Gini index, to choose predictors.^24^ We chose this method because it allowed us to select predictors based on their variable importance (i.e., association between predictor and response variable). Other methods such as stepwise regression rely on statistical significance and may result in inclusion of irrelevant predictors when used with large datasets. Because our composite outcome was rare, we used an under-sampled model to enhance the performance of our random forest.^25,26^ We combined related variables for both the random forest classification and the Poisson regression model because of small sample sizes for some of the variables. Food insecurity and housing; intimate partner violence and forced sex; all substance use; history of fetal and neonatal death; history of HDP and gestational diabetes; and history of cervical insufficiency or second trimester loss were combined into single variables. We included variables identifying diabetes, hypertension, and mental illness individually, and grouped all other chronic diseases into a single variable: (asthma, HIV, seizure disorder, renal disease, systemic lupus erythematosus). We categorized maternal race/ethnicity as Non-Hispanic White, Non-Hispanic Black, Hispanic, Non-Hispanic American Indian/Alaska Native, and because of small sample sizes, combined categories for Non-Hispanic Asian, Pacific Islander, Mixed Race, and Other Race. Variables with missingness were assigned a “missing” indicator so these observations were retained in the random forest classifier, though this indicator was not used in subsequent regression analysis.

We selected the top 15 variables (approximately top half) ranked by variable importance for Poisson regression using the full dataset. We used a complete-case analysis to model our composite outcome of adverse maternal events at birth and six weeks postpartum and present risk ratios and 95% confidence intervals (CIs).

This research was reviewed and approved by the University of North Carolina Office of Human Research Ethics (IRB #20-2241, initial approval date November 10, 2020).

## RESULTS

A total of 205,916 births to Medicaid beneficiaries met our inclusion criteria. In our sample, 6,156 (3.0%) had an adverse maternal event occurring between birth and up to six weeks postpartum, including 1,222 with SMM at birth (0.6%), 1,796 with ICU admission at birth (0.9%), and 3,635 with a prolonged birth hospitalization or postpartum readmission (1.8%). The overlap between these conditions is summarized in Figure 1. Most of our sample were aged 20-24 years (35.4%), had completed high school (38.1%), and had preexisting traditional Medicaid (53%; Table 1). Our sample was racially diverse, with 43.1% identifying as Non-Hispanic White, 37.9% as Non-Hispanic Black, 10.3% Hispanic, 2.4% as Non-Hispanic American/ Indian Alaska Native, and 6.3% Non-Hispanic Asian, Pacific Islander, Mixed, or Other.

**Figure 1.**
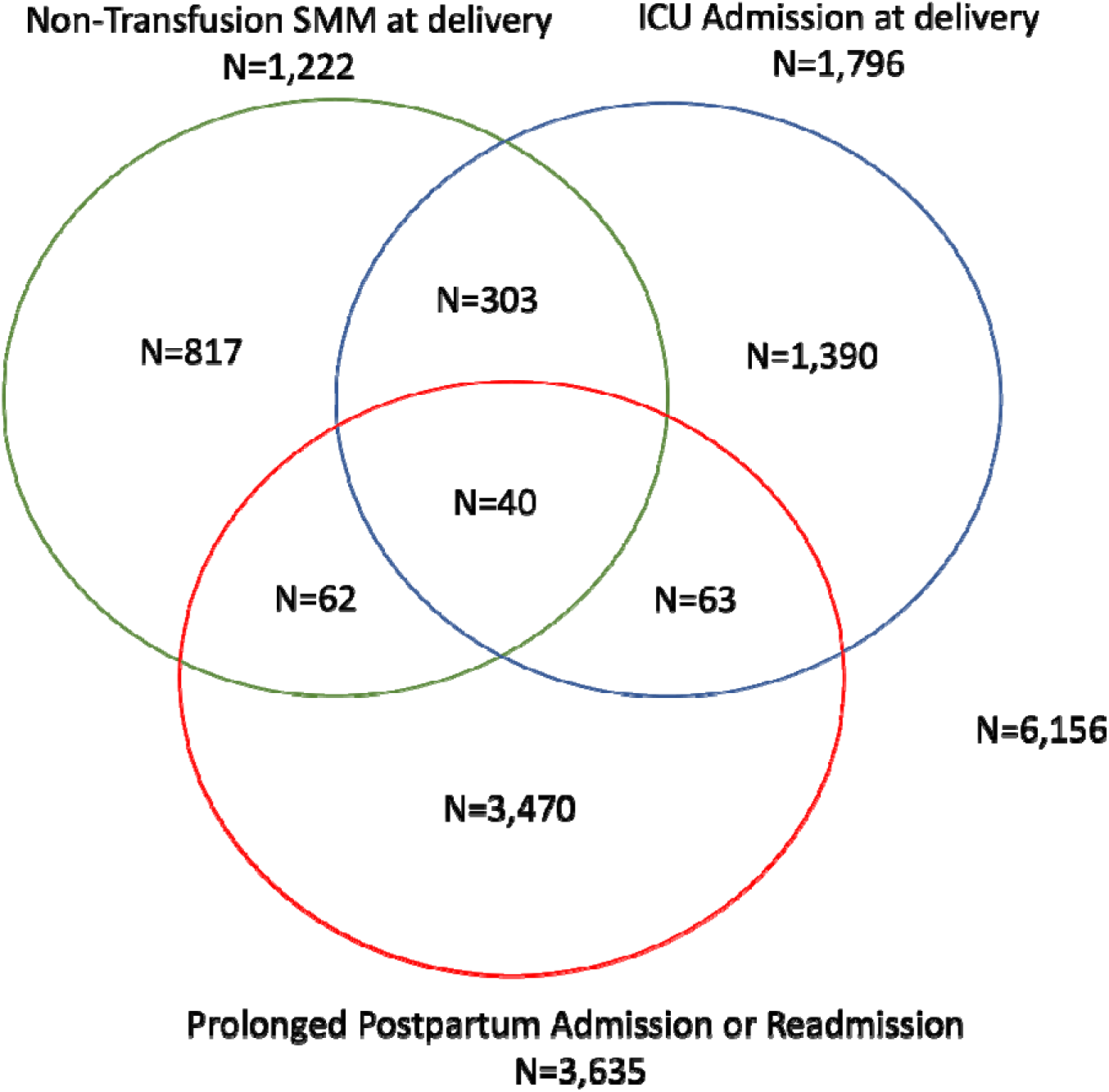
Adverse Maternal Events Among Medicaid Beneficiaries in the North Carolina Pregnancy Medical Home Program (2014-2019)

**Table 1.**
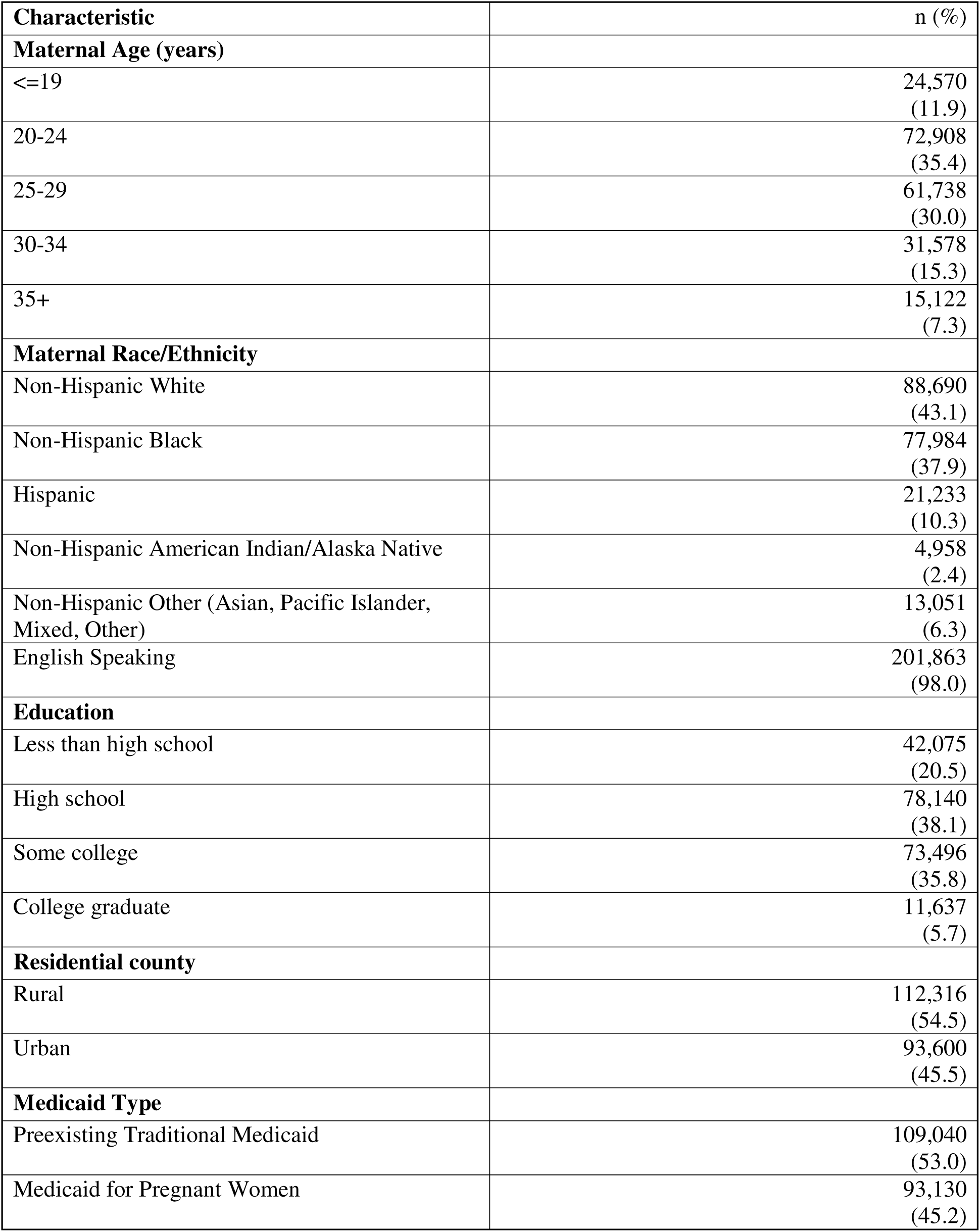
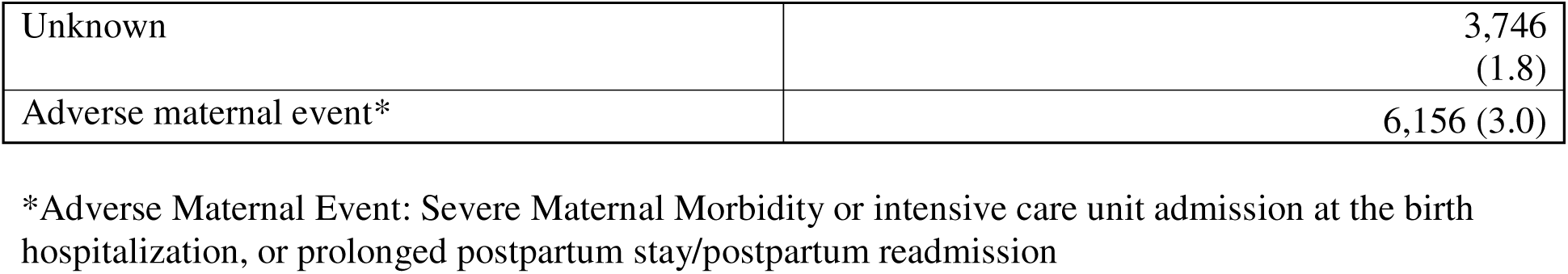
Characteristics of Medicaid Beneficiaries in the North Carolina Pregnancy Medical Home Program with a Live Birth from 2014-2019 (N=205,916)

| **Characteristic** | **n (%)** |
| --- | --- |
| **Maternal Age (years)** |  |
| <=19 | 24,570<br>(11.9) |
| 20-24 | 72,908<br>(35.4) |
| 25-29 | 61,738<br>(30.0) |
| 30-34 | 31,578<br>(15.3) |
| 35+ | 15,122<br>(7.3) |
| **Maternal Race/Ethnicity** |  |
| Non-Hispanic White | 88,690<br>(43.1) |
| Non-Hispanic Black | 77,984<br>(37.9) |
| Hispanic | 21,233<br>(10.3) |
| Non-Hispanic American Indian/Alaska Native | 4,958<br>(2.4) |
| Non-Hispanic Other (Asian, Pacific Islander, Mixed, Other) | 13,051<br>(6.3) |
| English Speaking | 201,863<br>(98.0) |
| **Education** |  |
| Less than high school | 42,075<br>(20.5) |
| High school | 78,140<br>(38.1) |
| Some college | 73,496<br>(35.8) |
| College graduate | 11,637<br>(5.7) |
| **Residential county** |  |
| Rural | 112,316<br>(54.5) |
| Urban | 93,600<br>(45.5) |
| **Medicaid Type** |  |
| Preexisting Traditional Medicaid | 109,040<br>(53.0) |
| Medicaid for Pregnant Women | 93,130<br>(45.2) |
| Unknown | 3,746<br>(1.8) |
| Adverse maternal event* | 6,156 (3.0) |

\*Adverse Maternal Event: Severe Maternal Morbidity or intensive care unit admission at the birth hospitalization, or prolonged postpartum stay/postpartum readmission

Figure 2 summarizes the relative variable importance of each predictor in the model. The top predictors that were chosen for Poisson regression were: maternal age, BMI, race/ethnicity, education, pregnancy intendedness, Medicaid type, rural residency, any substance use, chronic diseases, parity, late entry into prenatal care, smoking status, history of HDP, history of preterm birth, and food insecurity/housing instability. The risk of adverse maternal events by these selected characteristics is available in Table 2. Characteristics that were not selected for the prediction model are available in Supplemental Table 1. Those who were older, Non-Hispanic Black, lived in an urban area and had food insecurity or unstable housing were more likely to have an adverse maternal event (Table 2). Those with obesity, or chronic conditions (diabetes or hypertension) were also more likely to have an adverse maternal event. Many of the pregnancy history factors were also associated with adverse maternal events, including previous fetal or neonatal death, and having a history of pregnancy conditions such as previous preterm birth or HDP or gestational diabetes.

**Figure 2.**
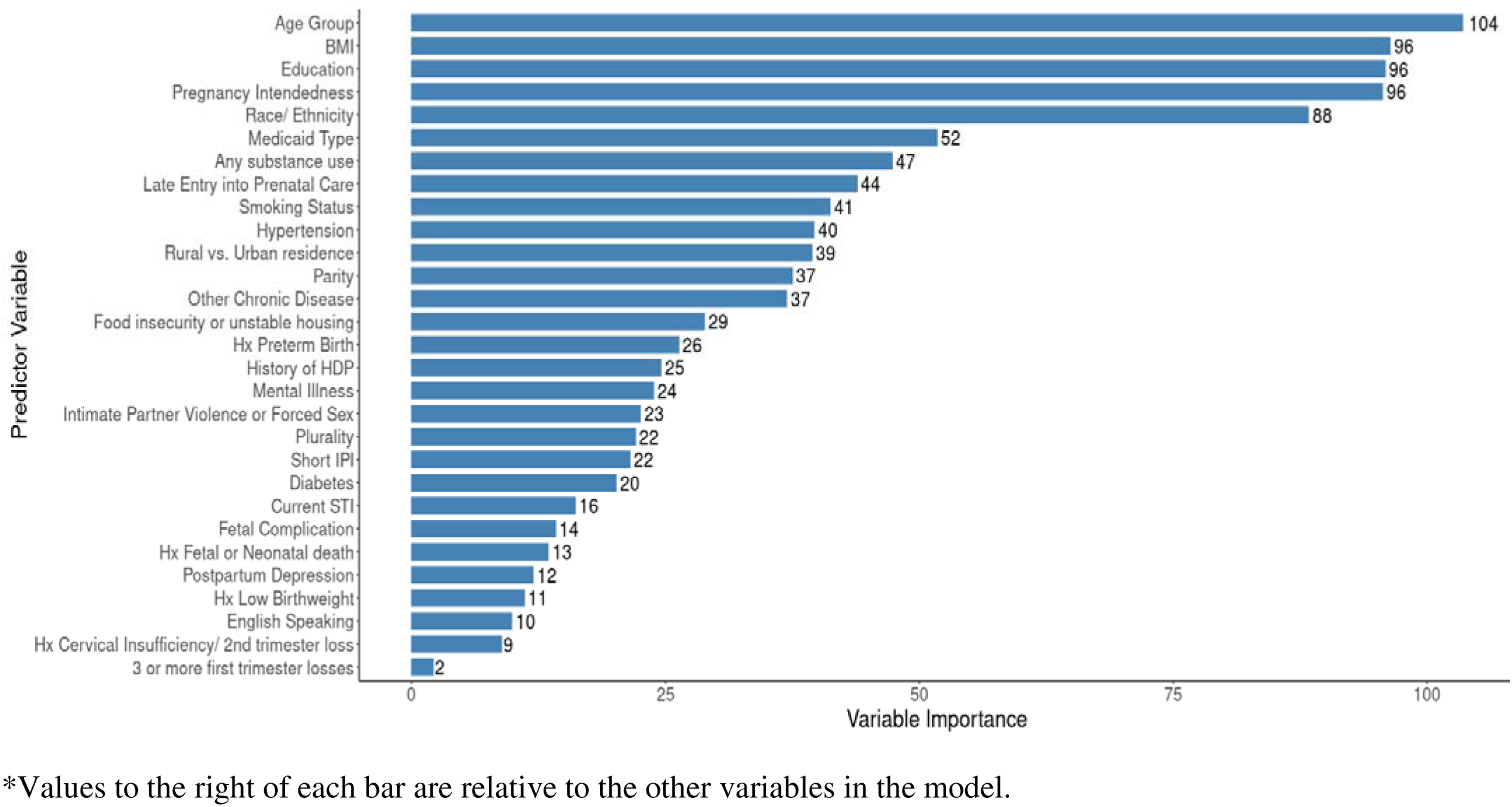
Variable Importance based upon the Gini Index in Random Forest Modeling*.

**Table 2:**
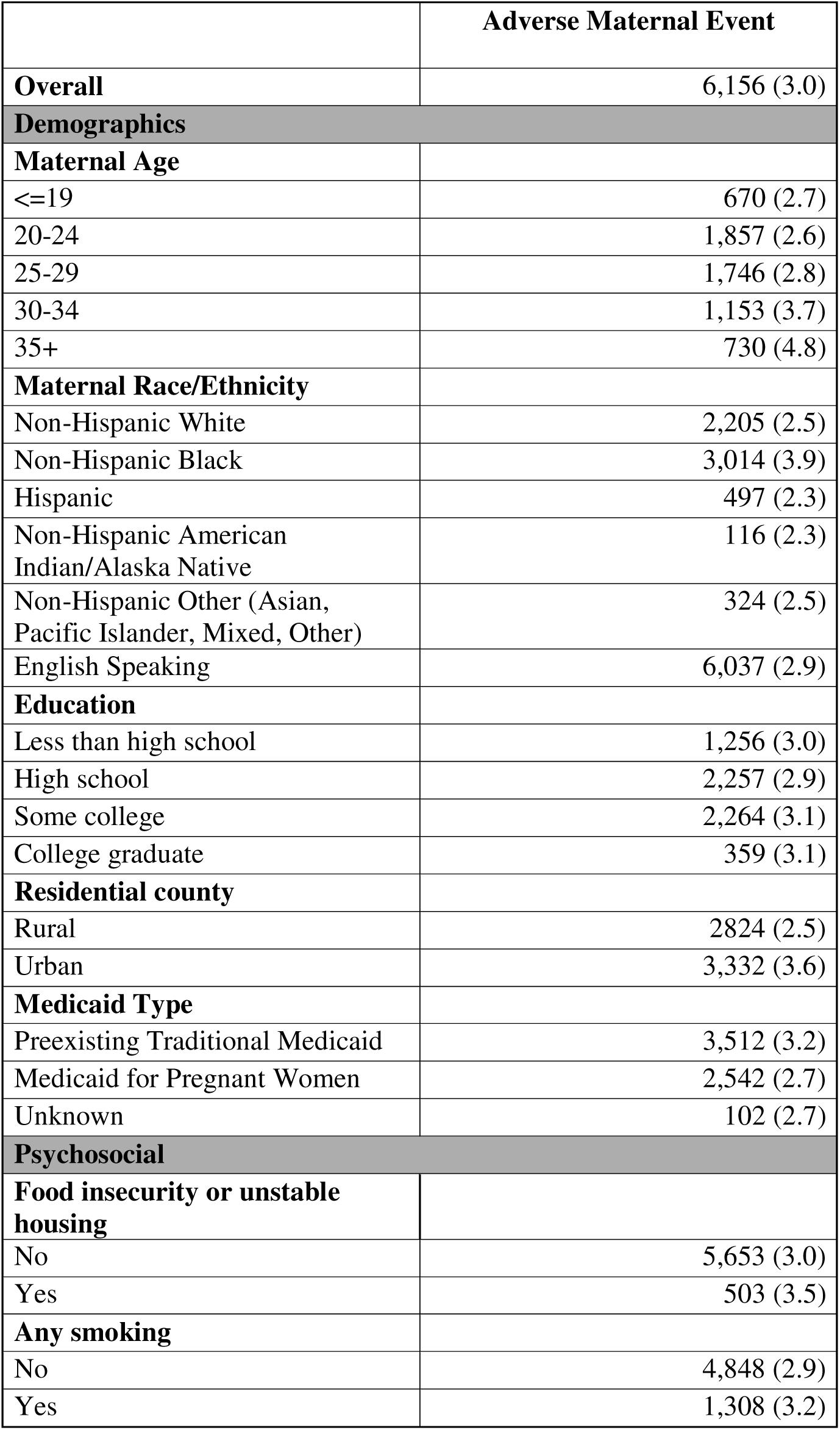
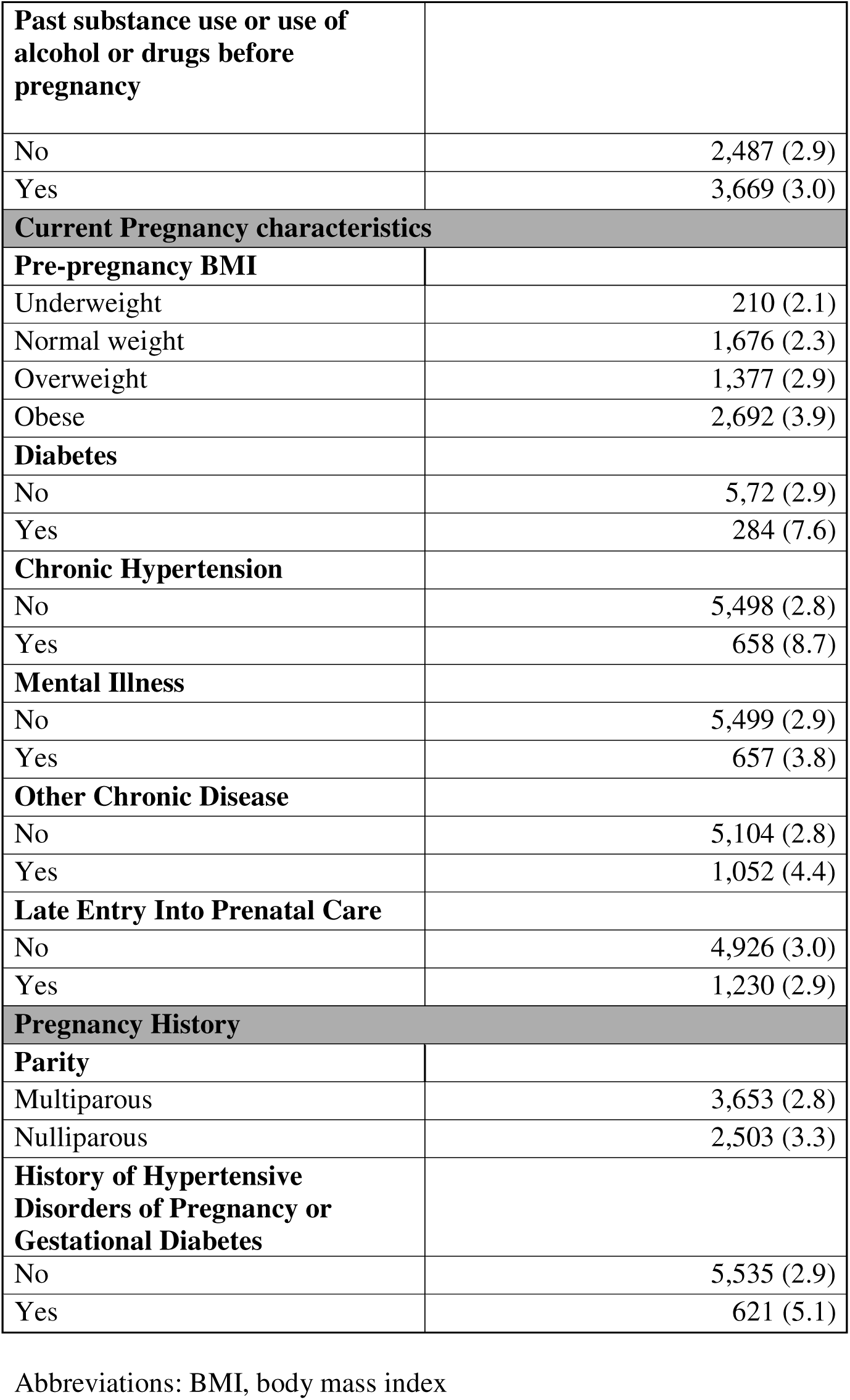
Risk of adverse maternal event by selected screening characteristics among Medicaid beneficiaries in the North Carolina Pregnancy Medical Home program with a live birth from 2014- 2019 (n=205,916)

The results of our Poisson model are shown in Figure 3. Although variables were selected based on their relative importance in the random forest model, not all variables were associated with SMM in the multivariable model. The Area Under the Curve (AUC) of our final model was 0.63 and at the point that both sensitivity and specificity were both optimized (predicted probability 0.032), sensitivity was 56% and specificity was 63%, indicating that our model had generally poor ability to discriminate between cases and non-cases. Maternal age > 25 years, smoking, BMI >25, diabetes, chronic hypertension, mental illness, other chronic diseases, nulliparity, history of preterm birth, and history of HDP or gestational diabetes were all associated with increased risk of adverse maternal events (Figure 3). Non-Hispanic Black pregnant people had increased risk of the composite outcome compared to Non-Hispanic White pregnant people, but Hispanic and Non-Hispanic AI/AN did not have elevated risk. Rural residence, having Medicaid for Pregnant Women, and late entry into prenatal care were associated with decreased risk of adverse maternal events. Increasing levels of education, pregnancy intendedness, food insecurity or housing instability, and any substance use were not significantly associated with risk of adverse maternal events and had risk ratios near the null.

**Figure 3.**
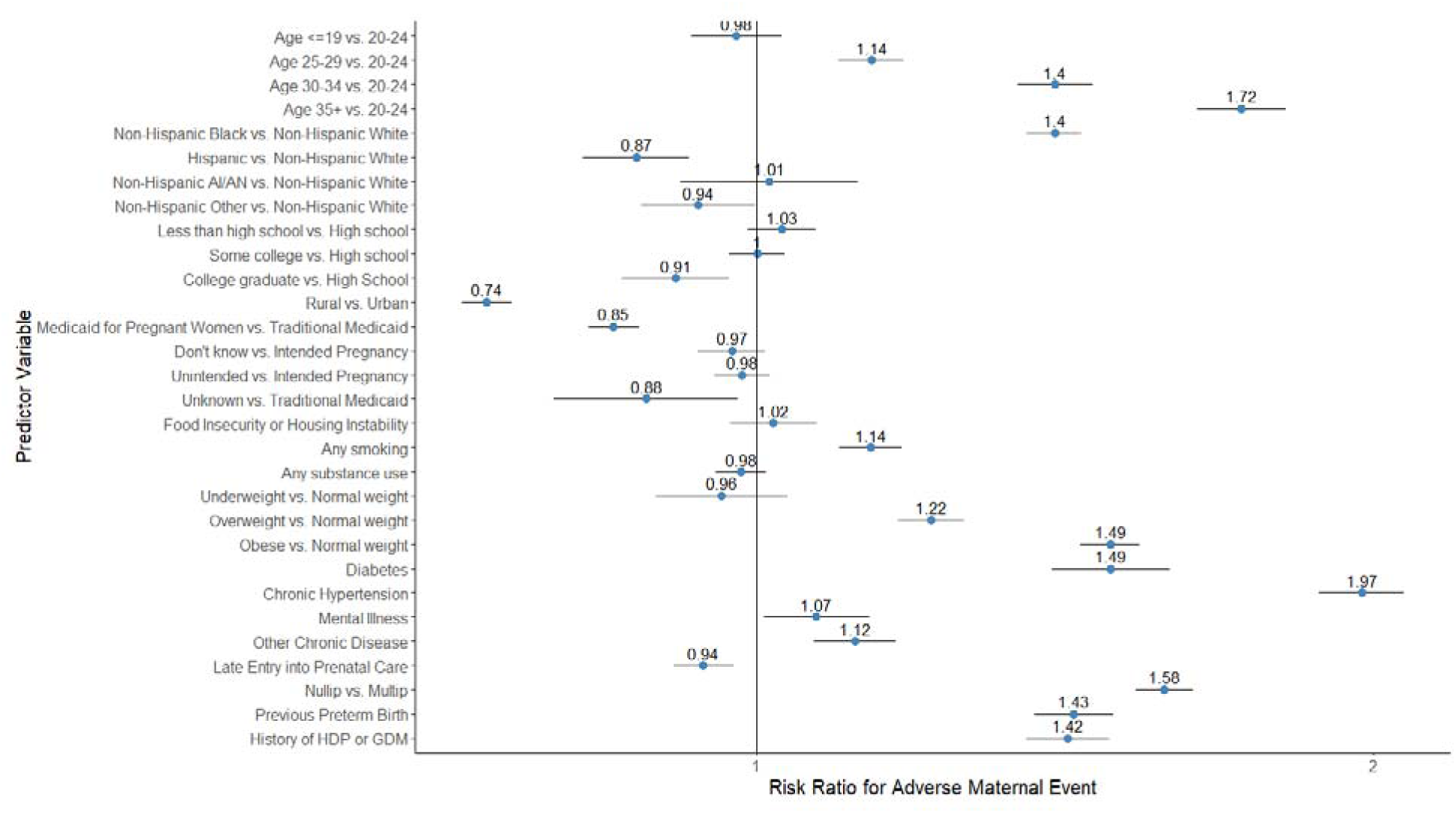
Risk Ratio (95% CI) of Adverse Maternal Events by Selected Prenatal Risk Screen Characteristics Using Poisson Regression.

## COMMENT

### Principal Findings

We found that the North Carolina PMH risk screen, designed to identify pregnant people at high risk of preterm birth, had factors that were associated with increased risk of adverse maternal events. These factors included maternal age greater than 25, Black race (as compared to White race), a BMI greater than 25, chronic diseases, smoking, nulliparity, and pregnancy history characteristics, such as a history of preterm birth or HDP. However, when these factors and selected elements of the risk screen were considered together in a model, they were poorly predictive of this composite outcome, which included SMM, ICU admissions, and postpartum hospitalizations.

### Results in the Context of What is Known

The North Carolina PMH was designed to reduce rates of preterm birth and low birth weight within the state by providing risk stratification and care management. While these efforts were successful in providing targeted care and in reducing these adverse neonatal outcomes, the association with maternal health outcomes has not been reported. In obstetric literature, it has been well documented that there is considerable overlap amongst those at risk of poor maternal health outcomes and those at risk of poor neonatal outcomes.^27–29^ It would stand to reason, therefore, that reductions in low birth weight and preterm birth because of targeted care management interventions might also address maternal clinical and social vulnerabilities that portend poor maternal health and obstetric outcomes.

A previous study by Tucker et al. used the same PMH risk screen data and population but with births from 2011-2012 to examine associations with preterm birth.^14^ Although we used different model selection methods, our final prediction model for adverse maternal events had substantial overlap with their model which predicted preterm birth. Differences between our models may be the result of different etiologies for infant and maternal outcomes or differences in our variable selection methods. For instance, factors such as history of cervical insufficiency, which has very low prevalence, may not contribute substantially to a maternal outcome model compared to an infant model. Tucker et al.’s final prediction model had a slightly higher degree of discernment, with an AUC of 0.66 compared to the 0.63 in our model. At the point that both were optimized, they reported lower sensitivity (44% vs 56%) but higher specificity (81% vs 63%).

Our study demonstrates that the current risk stratification model to identify those at risk of neonatal health outcomes was not similarly predictive of maternal health outcomes. There are likely several reasons why this discrepancy exists. First, our model’s poor prediction can reflect limitations in the design and implementation of the risk screen. For instance, it is possible that maternal risk factors that predict neonatal outcomes like LBW or PTB might be better captured on the risk screen while others that are important to the predictive modeling of maternal health outcomes are missing (e.g., prenatal care, other health care utilization), were incompletely filled out, or were not captured due to the timing at which the form was completed in pregnancy (e.g., preeclampsia, gestational diabetes). Second, the combined risk factors that we placed into the model are potentially poorly predictive due to collinearity as several of these risk factors are also associated with one another. Third, the maternal outcome of interest was a composite of SMM, ICU admissions, prolonged hospitalization and postpartum readmission. It is possible that components of this composite are less clearly associated with risk factors on the risk screening form either due to their rarity of occurrence (ICU admission) or the wide variation of clinical and social scenarios in which they might occur (prolonged hospitalization and readmission). Importantly, while the risk screen form is not associated with adverse maternal events as defined by our study team, it does not preclude a possible association with other maternal health outcomes that might be more closely associated with factors on the risk screen (e.g., health care utilization, postpartum mental health or mood disorders, HDP).

### Clinical Implications

This study is important for demonstrating the utility and limitations of programmatic risk stratification for reducing adverse maternal events. The role of patient-centered medical homes as well as risk assessment with care management have been well-documented within medical literature.^30^ Medical home models are heterogenous and complex, yet all are intended to provide comprehensive care to an individual through accessible, multi-disciplinary efforts present in both clinical and non-clinical settings.^30,31^ While early research in medical home models and care management was within both pediatrics and primary care, there is emerging evidence within obstetric care on their role in influencing clinical and procedural outcomes.^32,33^ However, there are few studies on the efficacy or predictive potential of risk stratification methods used by these programs and, to the best of our knowledge, none that look at the ability of these models to identify patients at risk for adverse maternal outcomes. We believe this study highlights the importance of both validating risk stratification systems to address the appropriate outcomes of interest and encouraging programs to consider balancing both maternal and neonatal outcomes. Further, the intervention for those who screen high risk is care management. There are analyses that have demonstrated that care management can influence neonatal outcomes, such as preterm birth or low birth weight, in North Carolina, South Carolina, and New York.^15,18,19^ While such an evaluation to assess the impact of care management on maternal outcomes was outside the scope of this analysis, we believe it is important to understand whether care management provided to the pregnant person can have direct effects on their health and clinical outcomes.

### Strengths and Limitations

One of the main strengths of this analysis was the use of a composite measure to capture adverse maternal events. In doing so, we include SMM as an important outcome but also broadened our perspective to other maternal health outcomes that have salience for patients, providers, and health systems, including those occurring after the labor and delivery period. Our sample was large and diverse, and we utilized rich data from the PMH risk screen to identify risk factors for our outcome, including social factors that would not have been possible to explore using live birth records or Medicaid claims alone. We also provide information from a state with an established PMH program for Medicaid beneficiaries, including an associated risk screen form, which can be used to inform similar programs being implemented in other states.

There are also several limitations of our study. The timing of data collection across data sources varied: information from the live birth record was collected at the time of birth, while risk screen forms were completed during a prenatal visit and prior to birth. Issues with temporality or mechanism of data collection might influence the accuracy of several variables within the study either because some factors might change over the course of a pregnancy (e.g., HDP) or because some data on the risk screen form might be more reliably captured on the birth certificate (and vice versa). To the best of our knowledge, there is no current validation study of the North Carolina PMH risk screen form with variables that are also collected on the birth certificate. Relatedly, because these are three large linked administrative datasets, there is the possibility for missing data and misclassification. As described in our methods section, there are limitations to using data that were not collected for research purposes. Future work in our program and others may have to consider how to best capture risk screen information to avoid such problems. However, this study was not intended to be an epidemiologic study of risk factors for SMM, ICU admission, and postpartum hospitalizations; rather this was a pragmatic assessment of the PMH risk screen form. Moreover, this study cannot account for variations in how the risk screen is implemented within clinic settings, how it is presented to a patient, or how one chooses to complete the form – variations in clinical practice, the manner in which the risk form is presented, or the patient’s level of trust in the health system might influence how individuals answer sensitive items on the form or whether they elect to participate in the PMH. For instance, people might underreport certain aspects of the risk screen such as their social history or substance use or might not complete the entire form thus limiting the sensitivity of this tool. Importantly, since the time period when these data were collected, the PMH has transitioned management as a result of NC Medicaid shifting to managed care and therefore, implementation might be increasingly variable. Last, because this program was ongoing, it is possible that those who received the intervention (care management) were able to avoid adverse maternal events, which would have changed our model’s predictive ability.

### Conclusions

Care management during pregnancy is a vitally important and relevant topic in the United States. As both public and private payers are increasingly incentivized to design programs that optimize maternal and fetal outcomes, it is important to consider how initiatives, like the North Carolina PMH can serve as a model and what lessons can be learned. While the North Carolina PMH demonstrates an effective risk screening model for the prediction of preterm birth, these same tools and their implementation were not designed and are not effective in predicting adverse maternal health outcomes. It is, therefore, important to consider how such programs can be designed to center both the mother and neonate so that both members of the dyad benefit. These considerations might include how risk stratification is constructed and validated, how best to strengthen systems and processes of risk stratification, what activities care coordination programs utilize, and what metrics are tracked and considered important. Aside from studying how these programs are designed is the equally crucial consideration of how programs are implemented. This study was a pragmatic study of a risk screen yet did not delve into how providers communicate the risk screen, how individuals answer questions or complete the form, or what happens once care management needs are identified. Future studies should expand upon our work to elucidate how best to identify those at risk of adverse maternal outcomes within related programs and, ultimately, how best to respond to the needs of clinically and socially vulnerable populations.

## Data Availability

Researchers with human subjects training can request the data from North Carolina Medicaid.

## Notes

### Competing Interest Statement

The authors have declared no competing interest.

### Author Declarations

The ethics committee of University of North Carolina gave ethical approval for this week.

### Funding

- NC Department of Health and Human Services: #30-2019-020-DHB

