## Supplemental Table 1 for "Risk Screening in a Medicaid-Managed Pregnancy Medical Home: The Need to Center Maternal Health Outcomes in Public Health Programming"

**Supplemental Figure 1: North Carolina Pregnancy Medical Home Risk Screen Form (version date: December 1, 2020)**

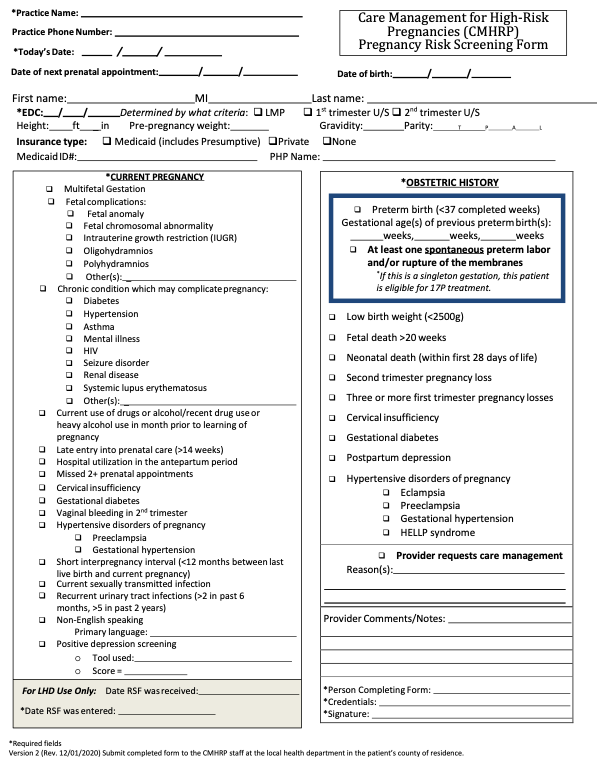

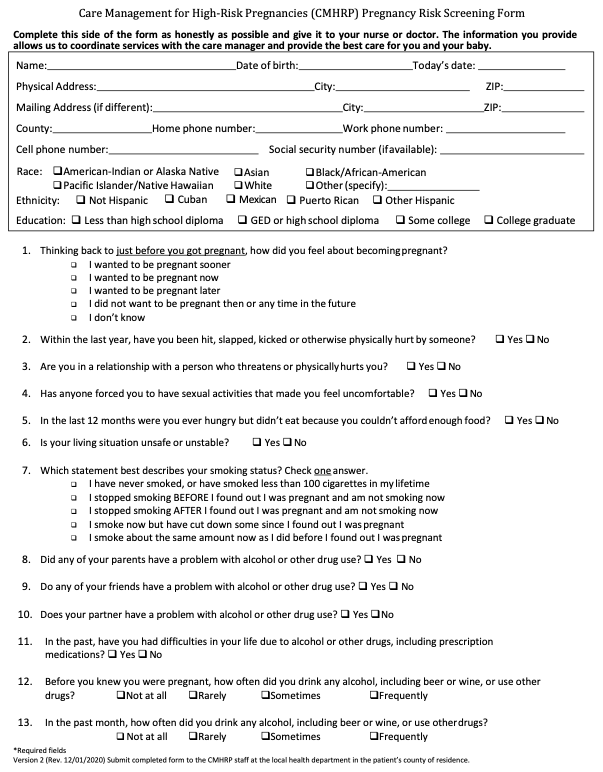
