## Supplemental Table 1 for "Risk Screening in a Medicaid-Managed Pregnancy Medical Home: The Need to Center Maternal Health Outcomes in Public Health Programming"

**Supplemental Table 1: Risk of Adverse Maternal Events by all Characteristics**

|  | Adverse Maternal Event |
| --- | --- |
| **Overall** | 6,156 (3.0) |
| **Maternal Age** |  |
| <=19 | 670 (2.7) |
| 20-24 | 1,857 (2.6) |
| 25-29 | 1,746 (2.8) |
| 30-34 | 1,153 (3.7) |
| 35+ | 730 (4.8) |
| **Maternal Race/Ethnicity** |  |
| Non-Hispanic White | 2,205 (2.5) |
| Non-Hispanic Black | 3,014 (3.9) |
| Hispanic | 497 (2.3) |
| Non-Hispanic American Indian/Alaska Native | 116 (2.3) |
| Non-Hispanic Other (Asian, Pacific Islander, Mixed, Other) | 324 (2.5) |
| **English Speaking** | 6,037 (2.9) |
| **Education** |  |
| Less than high school | 1,256 (3.0) |
| High school | 2,257 (2.9) |
| Some college | 2,264 (3.1) |
| College graduate | 359 (3.1) |
| **Residential county** |  |
| Rural | 2824 (2.5) |
| Urban | 3,332 (3.6) |
| **Medicaid Type** |  |
| Preexisting Traditional Medicaid | 3,512 (3.2) |
| Medicaid for Pregnant Women | 2,542 (2.7) |
| Unknown | 102 (2.7) |
| **Psychosocial Characteristics** | |
| **Pregnancy Intendedness** |  |
| Intended | 1900 (2.9) |
| Unintended | 2783 (2.8) |
| Don't know | 1,304 (3.2) |
| **Intimate Partner Violence or Forced Sex** |  |
| No | 5, 839 (3.0) |
| Yes | 317 (3.6) |
| **Food insecurity or unstable housing** |  |
| No | 5,653 (3.0) |
| Yes | 503 (3.5) |
| **Any smoking** |  |
| No | 4,848 (2.9) |
| Yes | 1,308 (3.2) |
| **Substance use in a partner, family member or friend** |  |
| No | 4,833 (3.0) |
| Yes | 1,323 (3.2) |
| **Past substance use or use of alcohol or drugs before pregnancy** |  |
| No | 2,487 (2.9) |
| Yes | 3,669 (3.0) |
| **Current Pregnancy characteristics** | |
| **Multiple gestation** |  |
| No | 5,910 (2.9) |
| Yes | 248 (7.4) |
| **Pre-pregnancy BMI** |  |
| Underweight | 210 (2.1) |
| Normal weight | 1,676 (2.3) |
| Overweight | 1,377 (2.9) |
| Obese | 2,692 (3.9) |
| **Has Fetal Complication** |  |
| No | 6,030 (3.0) |
| Yes | 126 (4.6) |
| **Diabetes** |  |
| No | 5,72 (2.9) |
| Yes | 284 (7.6) |
| **Chronic Hypertension** |  |
| No | 5,498 (2.8) |
| Yes | 658 (8.7) |
| **Asthma** |  |
| No | 5,700 (2.9) |
| Yes | 456 (3.7) |
| **Mental Illness** |  |
| No | 5,499 (2.9) |
| Yes | 657 (3.8) |
| **HIV** |  |
| No | 6,137 (3.0) |
| Yes | 19 (7.3) |
| **Seizure Disorder** |  |
| No | 6,064 (3.0) |
| Yes | 92 (4.4) |
| **Renal Disease** |  |
| No | 6,128 (3.0) |
| Yes | 28 (7.1) |
| **Systemic Lupus Erythematosus** |  |
| No | 6,131 (3.0) |
| Yes | 25 (8.3) |
| **Other Chronic Disease** |  |
| No | 5,104 (2.8) |
| Yes | 1,052 (4.4) |
| **Late Entry Into Prenatal Care** |  |
| No | 4,926 (3.0) |
| Yes | 1,230 (2.9) |
| **Short Interpregnancy Interval** |  |
| No | 5,883 (3.0) |
| Yes | 273 (2.3) |
| **Current STI** |  |
| No | 6,004 (3.0) |
| Yes | 152 (3.2) |
| **Pregnancy History** | |
| **Parity** |  |
| Multiparous | 3,653 (2.8) |
| Nulliparous | 2,503 (3.3) |
| **Previous Preterm Birth** |  |
| No | 5,463 (2.9) |
| Yes | 693 (4.7) |
| **Previous Low birthweight** |  |
| No | 6,018 (3.0) |
| Yes | 138 (4.2) |
| **Previous Fetal Death or Neonatal Death** |  |
| No | 5,970 (3.0) |
| Yes | 186 (5.6) |
| **Previous cervical insufficiency or 2^nd^ trimester loss** |  |
| No | 6,027 (3.0) |
| Yes | 129 (5.0) |
| **Three or More First Trimester Losses** |  |
| No | 6,141 (3.0) |
| Yes | 15 (5.3) |
| **History of Postpartum Depression** |  |
| No | 6,027 (3.0) |
| Yes | 129 (3.6) |
| **History of Hypertensive Disorders of Pregnancy or Gestational Diabetes** |  |
| No | 5,535 (2.9) |
| Yes | 621 (5.1) |

Abbreviations: BMI, body mass index; HIV, human immunodeficiency virus; STI, sexually transmitted infection;
